# A Translational Model of Compulsive Alcohol Use in Humans

**DOI:** 10.1101/2025.05.21.25328094

**Authors:** Colette Delawalla, Lydia Zindel, Sarah Jung, Alexandra Cohen, Irwin Waldman, Hope Derricott, Rohan H. C. Palmer

## Abstract

Alcohol use disorder (AUD) is heterogenous, and criteria can be met through several expressions of harmful use (Watts et al., 2021) ranging from frequent binge drinking episodes and low-grade excessive consumption (e.g., 2 drinks every evening), to everyday use associated with neurobiological changes seen in active addiction. Precise understanding of use patterns and the cognitive and emotional experiences that drive these patterns are key in both a research context, where neurobiological, genetic, and personality factors may differ amongst use patterns (e.g., Delawalla et al., 2023) and in a clinical intervention context, where intervention for one use pattern may look very different from another (Nadkarni et al., 2022). We derived a four-factor model of compulsive alcohol use (CAU) in a representative sample of N = 2,004 Americans. The CAU model suggests the construct is comprised of *Intrusive Thoughts, Emotionality, Craving,* and *Loss of Control.* Further, we validated the emergent models against collateral measures to verify the structure and predictive value.

## A Translational Model of Compulsive Alcohol Use in Humans

Alcohol use disorder (AUD) is heterogenous, and criteria can be met through several expressions of harmful use ranging from frequent binge drinking episodes and low-grade excessive consumption (e.g., 2 drinks every evening), to everyday use associated with neurobiological changes seen in active addiction (Watts et al., 2021). Precise understanding of use patterns and the cognitive and emotional experiences that drive these patterns are key in both a research context, where neurobiological, genetic, and personality factors may differ amongst use patterns (e.g., Delawalla et al., 2023) and in a clinical intervention context, where intervention for one use pattern may look very different from another (Nadkarni et al., 2022). Indeed, recent research suggests that the use of blunt instruments, such as those assessing quantity and frequency of alcohol use and Diagnostic and Statistical Manual of Mental Disorders, Fifth Edition, Text Revision (DSM-5-TR; APA, 2013) criteria of AUD, have hindered attempts at translational theory-building work in these areas (Watts et al., 2021). Further, the use of these instruments is a particularly salient issue in “compulsive substance use”, which is reasonably well-defined in the preclinical literature but has yet to be translated meaningfully in humans.

### Compulsivity and Alcohol Use

If one considers *engagement* with alcohol (as a spectrum whereby the left side represents use that is not associated with any markers of dependence (such as craving), the middle represents an occasional desire for alcohol for adaptive purposes (e.g., social enmeshment, wine paring for an enhanced culinary experience) and the right extreme represents active addiction (e.g., craving, withdrawal, tolerance, loss of control), compulsive alcohol use would exist on the furthest right extreme. In this representation, *engagemen*t with alcohol captures the context and individual experience before, during, and after consumption and is not synonymous with quantity and frequency of use. In studies conducted with animals (e.g., model organisms such as rodents, mice, and non-human primates), henceforth referred to as preclinical, this construct is understood as one of the most entrenched expressions of addiction. Though definitions vary (see Robbins et al., 2024 for thorough review), common ground across theories suggest compulsivity in substance consumption 1) is a trait-based difference such that some individuals are more prone than others (Berridge et al., 1989; Berridge and Robinson, 2016), 2) is an automatic process whereby goal directed behavior is overcome by a very strong maladaptive habitual response (Luscher et al., 2020), 3) may be a response to interoceptive cues, such as negative affect or withdrawal (Koob and Le Moal, 2008) and 4) results when negative consequences fail to deter the behavior (Luscher et al., 2020).

Preclinical operationalizations of compulsivity align with broad clinical definitions of the construct. Specifically, compulsivity is a trait that may result in actions (overt or covert) made in response to a stimulus (internal or external) that are automatic, repetitive, and excessive, and interfere with an individual’s functioning such that engagement in the behavior is at odds with one’s goals (Robbins et al., 2024). Likewise, the literature on compulsivity acknowledges that compulsive behaviors do not exist in a vacuum—they appear within the context of emotions, cognitions, and urges. Indeed, this maps cleanly onto *conceptual* understandings of advanced alcohol addiction whereby an individual consumes the substance in response to a cue (e.g., withdrawal symptom, emotional experience), in an automatic, repetitive, and excessive manner even when the action does not align with their goals (e.g., going to work) and there are negative consequences. However, direct translations of this construct from preclinical rodent models to humans are few and far between.

One reason for the dearth of translation is that the DSM I, II, III, III-R IV, IV-TR, V, V-TR criteria for substance use disorders were developed independently from the empirical literature on addiction and exist for the purpose of identifying individuals in need of intervention, and not for the purpose of testing and expanding theory. As such, though the DSM hosts a spectrum in the form of “mild”, “moderate”, and “severe” AUD, these criteria fail to acknowledge a full range of use experiences outside of what can be defined as “disordered”. Possible combinations of DSM criteria that might capture compulsive use fall short as they do not acknowledge urges that drive use, the repetitive and/or excessive nature of the behavior, or associated cognitive and emotional experiences. Because of this, an individual could be diagnosed with a “severe” alcohol use disorder without exhibiting compulsivity, or by exhibiting compulsivity and only meet criteria for a “mild” or “moderate” use disorder, despite experiencing biological addiction. Thus, DSM criteria are not a reliable means of capturing compulsive alcohol use in humans.

### Translation Attempts of Compulsive Use

The preclinical literature is rich with several testable, and falsifiable operationalizations capturing compulsive substance use (CSU) in model organisms (e.g., Incentive-Salience theory [Berridge et al., 1989], Everitt and Robbin’s [2005] concept of “seeking”). However, to date, no such operationalization of CSU exists for humans. Likewise, there have been no direct translational attempts of these experimental paradigms in humans, largely because of ethical constraints. Further, when direct translational attempts have been made (e.g., Cyders et al., 2021) the methodologies used (e.g., intravenous alcohol administration) though valuable, are generally not scalable thus sample sizes are small.

Taking a latent variable approach to define compulsive alcohol use in larger samples of humans may provide a fruitful path forward. Indeed, this would facilitate review of the current literature to identify possible core components to submit to empirical testing in a scalable manner. Further, following Cronbach and Meehl’s (1955) method for establishing construct validity (i.e., establish internal structure, factor analyze, explore the nomological network) of compulsive alcohol use as a construct would provide a framework for testing the concept outside of experimental paradigms. This approach could establish an empirically derived phenotype that could be employed with greater precision to target the biological mechanisms of compulsive alcohol use in humans and eventually inform treatment.

## Current Study

The present study implemented Cronbach and Meehl’s (1959) construct validation procedure in two parts. The first was to develop a translational, empirically based model of compulsive alcohol use and second, to validate the derived model. These aims were completed within the same sample but consist of different measures and analytical approaches. As such, we first present participant information for each part of the study. Then we present the hypotheses, methods, and results of part one—development of the model—followed by the hypotheses, methods, and results of part two—validation of the selected model.

### Participants

Participants (N = 2,004) were recruited from Forthright Access (ForthrightAccess.com), an online, research recruitment panel. In order to participate in the study, individuals needed to be aged 22-45 and reported consuming a minimum of four alcohol beverages in the prior 30 days. The age range was carefully selected to account for the differential social norms and biological impacts across the lifespan. First, heavy drinking in emerging adulthood is common and *many* individuals “mature out” of heavy drinking patterns with the onset of more responsibility (Lee and Sher, 2018). In establishing a model of compulsive alcohol use, we wanted to reduce possible noise of younger drinkers who would go on to “mature out” in an effort to select individuals who either had a slower maturing out progression or who had failed to do so. Second, alcohol is metabolized less efficiently in older adults, leading to greater BAC levels after lower consumption, decreased cognitive function, particularly with regard to working memory, and greater sex differences in impairment as age increases (White et al., 2023). Given the presence and significance of age-related changes, we limited the age range to better capture “baseline” experience. Future directions will include testing the model in a full age range.

Additionally, we sought to include people who had exposure to alcohol with some degree of regularity. Recent epidemiological data suggests that on average Americans consume alcohol 1.69 days per week (Dawson et al., 2015). By setting the threshold for inclusion at four beverages in the prior 30 days, we were able to capture the low average frequency of use and/or significant drinking events that did not classify as a binge episode. In ensuring this degree of exposure we eliminated noise in analyses related to extremely low exposure consumers and enrich the sample to have a better chance at capturing compulsive alcohol users.

After meeting the above screening criteria, participants were invited to complete an informed consent process. Next, participants completed a battery of assessments capturing demographic information, personality, emotion, cognition surrounding alcohol consumption, day-to-day functioning, substance quantity and frequency, and alcohol related problems. In line with recommendations for ensuring participants provide quality data in online studies (e.g., no “Christmas tree” profiles) we included five attention check questions (e.g., “select option 3”; Newman et al., 2021) meant to identify persistent random responding. Failing three or more checks resulted in immediate removal from the study. Upon finishing the study, participants were paid $5—payment in line with the hourly minimum wage based on the average prorated study completion time. Demographic information can be found in Table 1. Participants were representative of the American population based on 2020 census data (U.S. Census Bureau, 2020). The sample represented all 50 states, approximately half were female (50.8%), and 58% were white, 21% Black, 15% Hispanic, and 5% Asian. Past year income was normally distributed and in line with census data. Likewise, 59.3% of the sample had at least an associate degree.

**Table 1.**
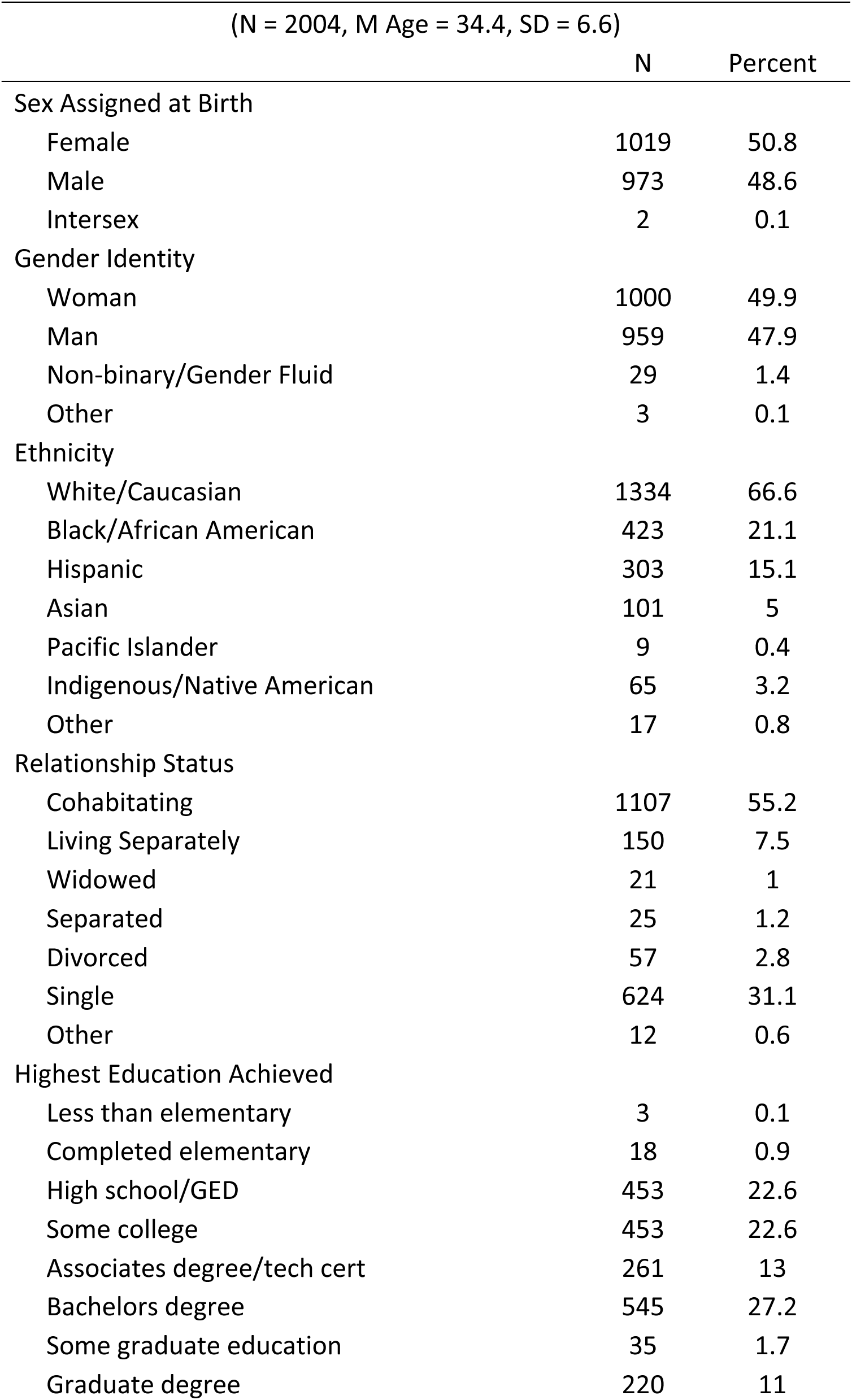
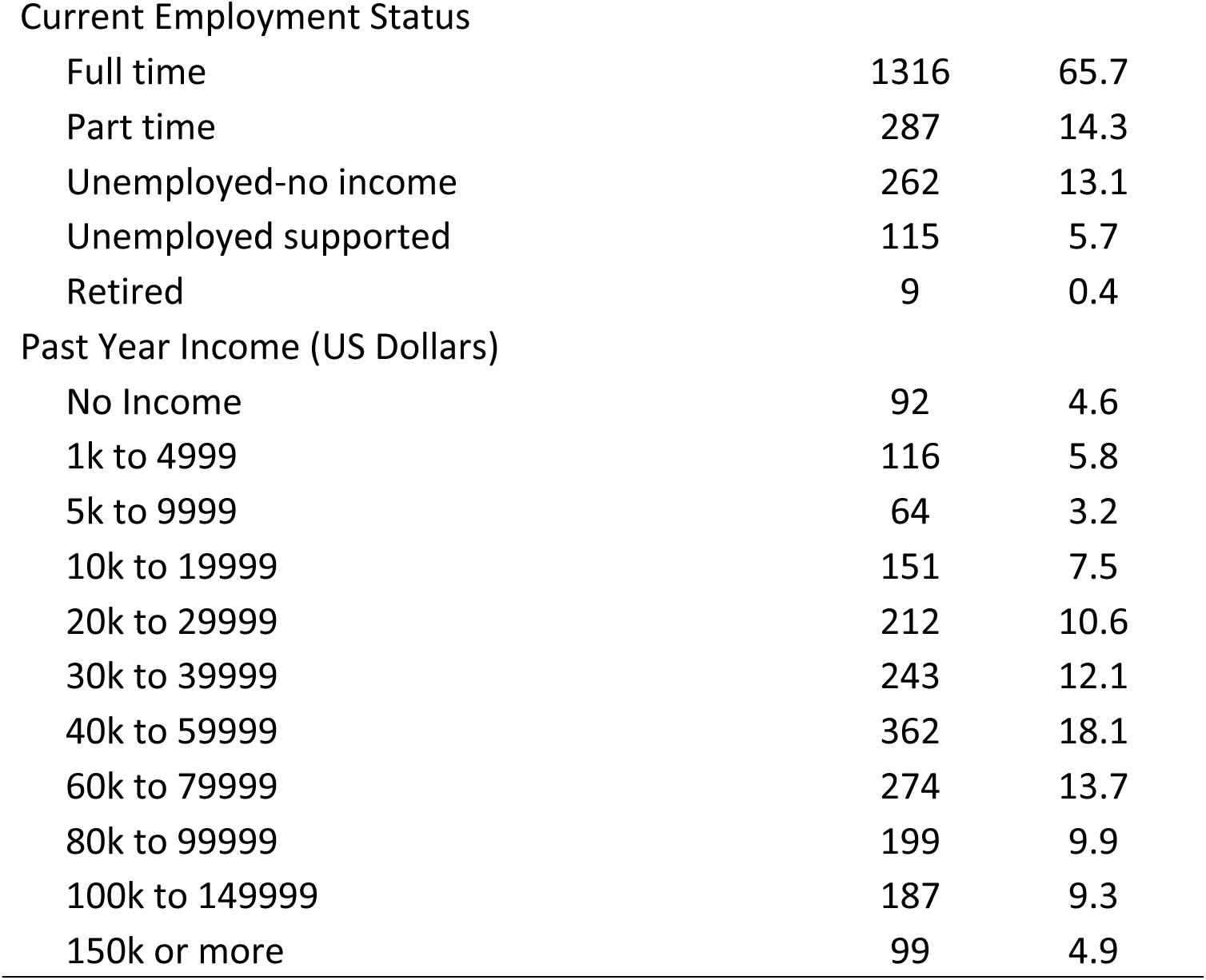
Participant Demographics.

| (N = 2004, M Age = 34.4, SD = 6.6) |  |  |
| --- | --- | --- |
|  | N | Percent |
| Sex Assigned at Birth |  |  |
| Female | 1019 | 50.8 |
| Male | 973 | 48.6 |
| Intersex | 2 | 0.1 |
| Gender Identity |  |  |
| Woman | 1000 | 49.9 |
| Man | 959 | 47.9 |
| Non-binary/Gender Fluid | 29 | 1.4 |
| Other | 3 | 0.1 |
| Ethnicity |  |  |
| White/Caucasian | 1334 | 66.6 |
| Black/African American | 423 | 21.1 |
| Hispanic | 303 | 15.1 |
| Asian | 101 | 5 |
| Pacific Islander | 9 | 0.4 |
| Indigenous/Native American | 65 | 3.2 |
| Other | 17 | 0.8 |
| Relationship Status |  |  |
| Cohabiting | 1107 | 55.2 |
| Living Separately | 150 | 7.5 |
| Widowed | 21 | 1 |
| Separated | 25 | 1.2 |
| Divorced | 57 | 2.8 |
| Single | 624 | 31.1 |
| Other | 12 | 0.6 |
| Highest Education Achieved |  |  |
| Less than elementary | 3 | 0.1 |
| Completed elementary | 18 | 0.9 |
| High school/GED | 453 | 22.6 |
| Some college | 453 | 22.6 |
| Associates degree/tech cert | 261 | 13 |
| Bachelors degree | 545 | 27.2 |
| Some graduate education | 35 | 1.7 |
| Graduate degree | 220 | 11 |
| Current Employment Status |  |  |
| Full time | 1316 | 65.7 |
| Part time | 287 | 14.3 |
| Unemployed-no income | 262 | 13.1 |
| Unemployed supported | 115 | 5.7 |
| Retired | 9 | 0.4 |
| Past Year Income (US Dollars) |  |  |
| No Income | 92 | 4.6 |
| 1k to 4999 | 116 | 5.8 |
| 5k to 9999 | 64 | 3.2 |
| 10k to 19999 | 151 | 7.5 |
| 20k to 29999 | 212 | 10.6 |
| 30k to 39999 | 243 | 12.1 |
| 40k to 59999 | 362 | 18.1 |
| 60k to 79999 | 274 | 13.7 |
| 80k to 99999 | 199 | 9.9 |
| 100k to 149999 | 187 | 9.3 |
| 150k or more | 99 | 4.9 |

## Study One: Development of Compulsive Alcohol Use Model

The aim of part one was to derive an empirical model of compulsive use in humans using exploratory and confirmatory factor analysis. Though exploratory in nature, we hypothesized that a model of compulsive alcohol use would emerge, consisting of factors that are theoretically relevant and empirically supported by the data.

### Methods

Assessment questions were derived from existing alcohol scales that captured aspects of what has been conceptualized as compulsive drinking. Measure included covered craving for alcohol, difficulty controlling one’s self around alcohol *and* difficulty stopping drinking once starting, emotional expectancies of drinking, obsessive thoughts, and thoughts and feelings of uncontrollable urges.

#### Measures

**Alcohol Urge Questionnaire (AUQ).** The Alcohol Urge Questionnaire (AUQ; Bohn et al., 1995) is an eight-item self-report measure assessing present desire to consume alcohol. Items include “I crave a drink right now” and “I do not need to have a drink right now” and are answered on a seven-point Likert scale from *Strongly Disagree* to *Strongly Agree*.

**Obsessive-Compulsive Drinking Scale (OCDS).** The Obsessive-Compulsive Drinking Scale (OCDS; Anton et al., 1995) is a widely used, 14-item questionnaire assessing the presence, intensity, and functional impact of alcohol related thoughts over the prior 30 days. Individuals completing the scale rate each item’s impact on them ranging from not at all (0) to extremely impactful (4). Importantly, the item response options are not consistent throughout the scale and each group of responses is tailored to the question. For example, ratings for the question “How frequently do these thoughts occur?” range from 0 *Never*, to 4, *Thoughts are too numerous to count and an hour rarely passes without several such thoughts occurring*. The scale yields a total score, a score for the Obsessive Subscale and a score for the Compulsive subscale. In this present study, we excluded items 7, 8, 9, and 10 as they assessed quantity and frequence of use and functioning—two variables we aimed to use in the validation portion of the study. Thus, we did not want to contaminate the item pool.

**Alcohol Craving Questionnaire, Short-Form Revised (ACQ-SFR).** The Alcohol Craving Questionnaire, Short-Form Revised (ACQ-SFR; Singleton and Gorelick, 1998) is a 12-item measure assessing current alcohol cravings and drinking expectancies. Items include “I want to drink so bad I can almost taste it” and “Drinking would put me in a better mood”, which participants rate on a seven-point Likert scale from *Strongly Disagree* to *Strongly Agree*. The ACQ-SRF was derived from the original 47-item ACQ, and the short form captures four dimensions of alcohol craving: compulsivity, expectancy, purposefulness, and emotionality.

**Impaired Control Scale - Part 3 (ICS).** The Impaired Control Scale (ICS, Pt 3; Heather et al., 1993) is a three-part scale assessing past and present attempts to limit drinking and the experiences that followed. For the proposed study we used Part 3, a 10-item section assessing current craving, participants’ beliefs about how they may react to attempting to limit alcohol, and current attitudes about drinking. Items include “I would have an irresistible urge to continue drinking once I started” and “I could slow down my drinking if I wanted to” and answer the items on a 5-point Likert scale from *strongly agree* to *strongly disagree*.

### Data Analysis

To explore possible models, the sample was randomly split into halves—a standard practice used to verify the emergent structure (Lorenze-Seva, 2022). In the first half, exploratory factor analysis (EFA) was conducted using item level data to understand the emergent factor structure. Standard procedures for understanding the ideal number of factors were applied (i.e., parallel analysis, VSS, Scree plot analysis), different factor rotation methods were used, and emergent models were ranked in terms of quality of fit, parsimony, and adherence to theory. In the second half, confirmatory factor analysis (CFA) was conducted based on the findings from the EFA in the first half of the sample. We tested and compared the fit of multiple alternative models to ensure we selected a model that represented the best balance of theory and model fit. Finally, a CFA was conducted in the full sample, to verify the structure of the data. Of note, due to overlap in items across measures, we expected a number of Heywood cases that would need to be addressed in this first stage.

### Results of Part One: Development of Compulsive Alcohol Use Model

**Exploratory Factor Analysis.** Item descriptives and inter-item correlations can be found in the Supplemental Materials Tables 1 and 2. In the random half, we evaluated the number of factors to retain using a Scree plot (that suggested 3-4 factors), Non-graphical Scree (that suggested 2-3 factors), Parallel Analysis (that suggested 11 factors with 5 components), Very Simple Structure (that suggested 1-2 factors), Maximum Likelihood (that suggested 4 factors), and Velicer MAP (that suggested 7 factors). Given the breadth of possible number of factors, we tested EFA’s of 2-7 factors using promax and geomin oblique rotation methods (all EFA factor loadings can be found in Supplemental Materials). Fit indices for each of the 12 models (2-7 factor promax and geomin EFA models) can be found in Table 2.

**Table 2.**
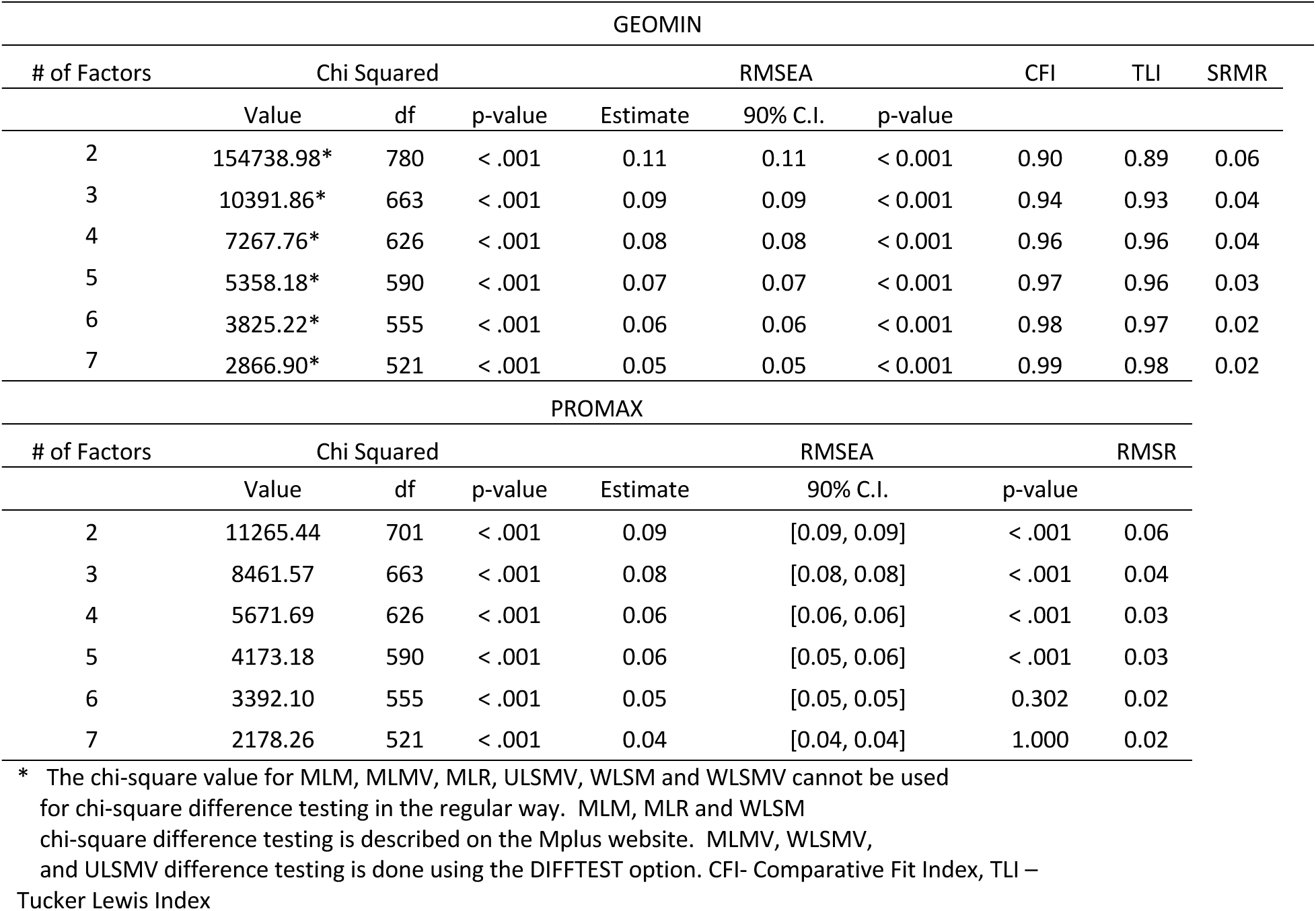
EFA Fit Indices.

In weighing model fit indices, parsimony of the emergent models, and factor loadings, we selected the three and four-factor models for comparison. Ultimately, we selected the four-factor promax rotated structure as the best solution. The four-factor promax rotated model had no cross-loaded items, had acceptable model fit (based on RMSEA < .08, CLI and TLI > .9; Hu and Bentler, 1999), and included the most interpretable and parsimonious solution with regard to item content and theory (Figure 1). Of the 40 items, 31 loaded onto a factor with a standardized loading ≥.4 (a commonly accepted cutoff point for loadings; Hu and Bentler, 1999). Notably, the nine items that failed to load onto any factors in this model consistently showed low factor loadings across other models (e.g., -.1 to .1).

**Figure 1.**
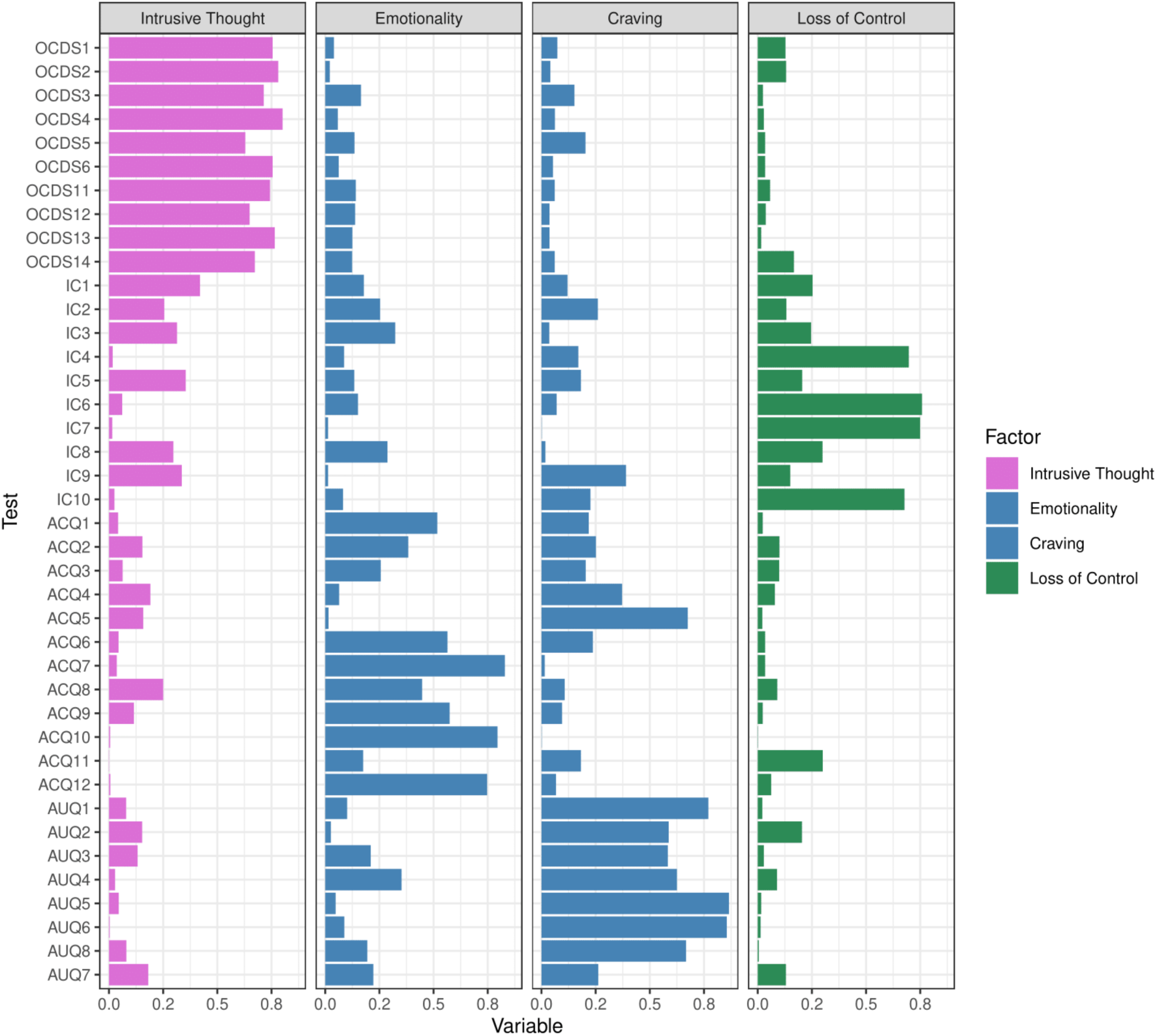
EFA Promax, Four Factor Model Factor Loadings. Note. All item loadings on four factor, promax in EFA model.

The selected model’s factors appear to represent *Intrusive Thoughts* about alcohol use, *Craving*, *Emotionality*, and *Loss of Control*. Comparison of the three and four-factor solutions show that in the three-factor solution a very large factor with craving and emotionality items splits in the four-factor model. This is important for two reasons. First, theory supports that emotionality is a key and separable aspect of addiction (Koob and LeMoal, 2008), thus an explicit factor capturing emotionality separate from craving is warranted. Second, in the four-factor model, each factor included a more suitable number of items (Factor 1 = 11 items, Factor 2 = 8 items, Factor 3 = 8 items, Factor 4 = 4 items) compared to the three-factor solutions. We also evaluated the five-factor models, these models excluded six items, and the fifth factor was a split of the loss of control factor without a clear demarcation in content.

**Confirmatory Factor Analysis.** We moved forward to the confirmatory factor analysis phase to test the following models in the second half of the sample: 1) a higher order dimension with four lower-order factors, 2) 4 correlated factors, 3) 3 correlated factors, and 4) items matched to scales. All models were based on the promax rotations and excluded items that failed to load ≥.4 in the EFA phase.

Fit indices for all CFA’s can be found in Table 3. Overall, the four-factor models showed marginally better fit than the three-factor models, which aligns with findings from the EFAs. The four factors were highly correlated in both models with estimates ranging from rs = .55-.86. This suggests the factors are distinct but related. Between the higher order and correlated factors model, the correlated factors model had a slightly better RMSEA. However, the higher order model was better aligned with theory suggesting that CSU is a combination of behaviors, cognitions, and emotional experiences occurring simultaneously, such that each component may be necessary but not sufficient on its own to capture the core construct. As such, we selected the higher order + four lower-order factor model comprising *Intrusive Thoughts, Craving, Emotionality,* and *Loss of Control* (Figure 2).

**Figure 2.**
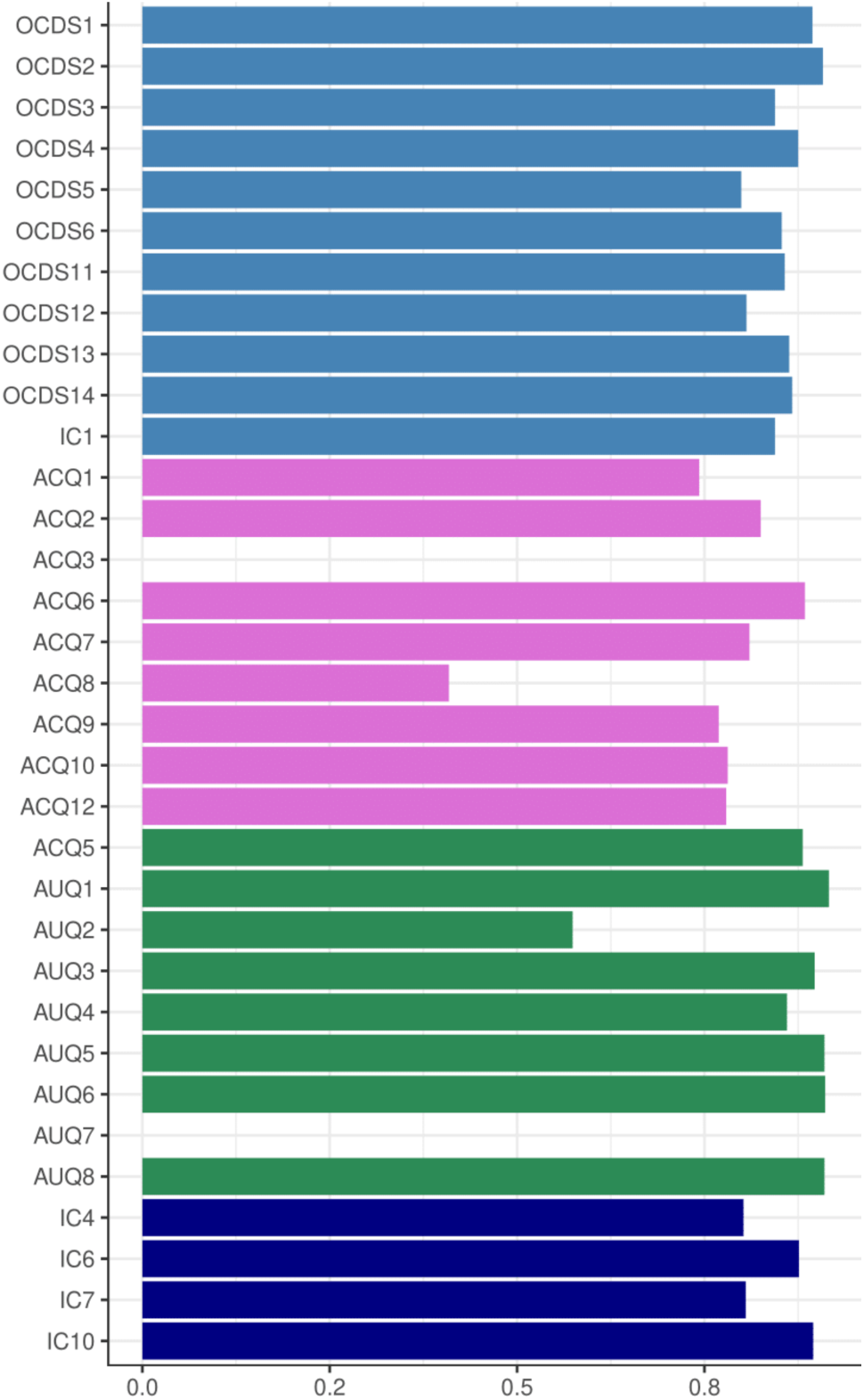
CFA Full Sample Selected Model Factor Loadings. Note. All item loadings in the full sample, for four factor higher order CFA model.

**Table 3.**
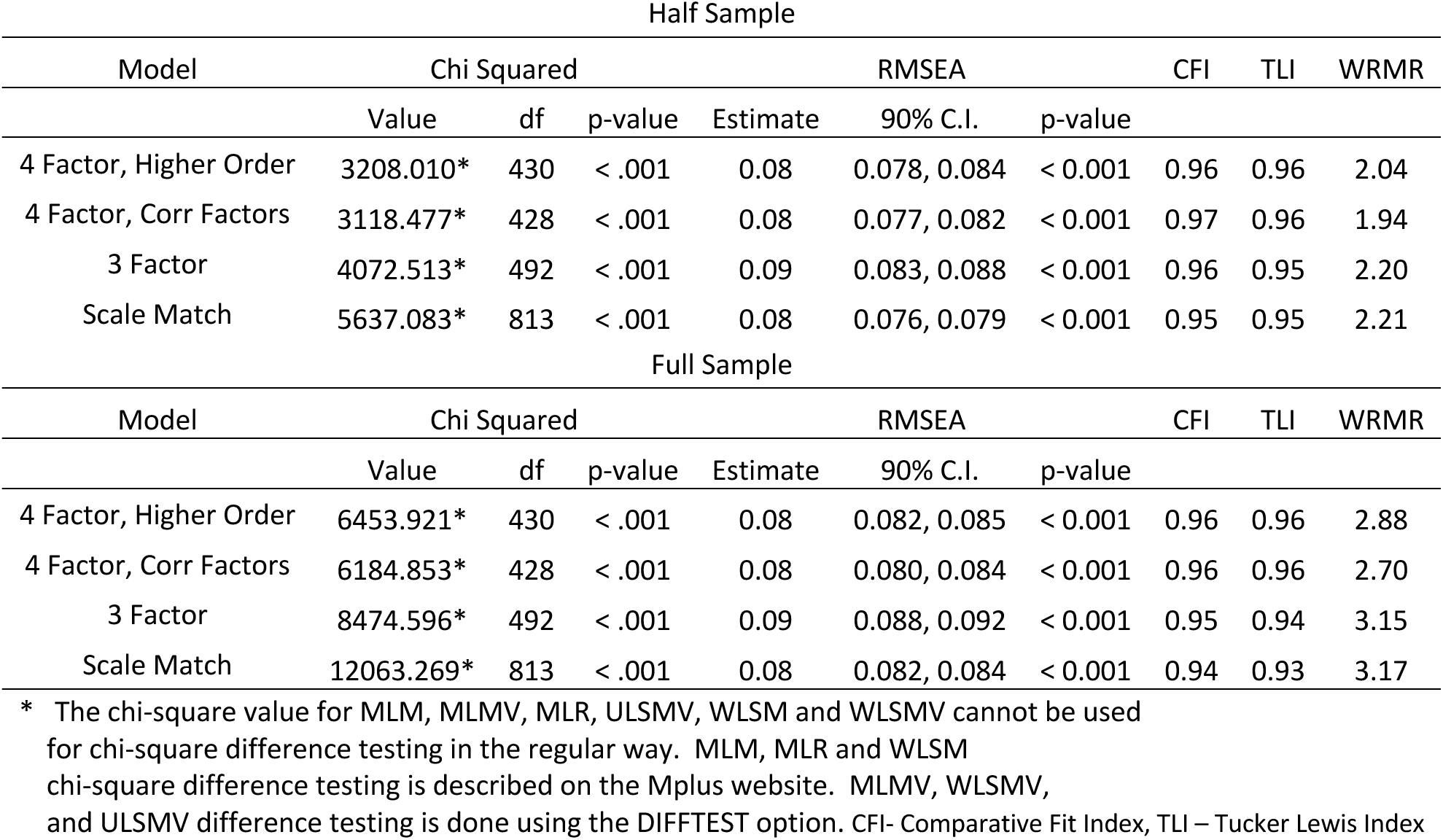
CFA Fit Indices.

As a final step in ensuring the consistency of the four-factor higher order model, we replicated the CFA in the entire sample. The model fit the data well (RMSEA= .08, TLI = .96, CLI = .96, loadings were all > 0.4). Model fit indices and factor loadings were consistent with those observed in the CFA in the second half of the data (Table 3).

## Part Two: Validation of Four-Factor Model of Compulsive Alcohol Use

The aim of part two was to validate the selected model of compulsive alcohol use— specifically, the four-factor higher order model chosen in part one. To ensure each factor was sufficiently distinct from the others, we investigated differential associations across the factors with collateral measures assessing trait and state impulsivity, state emotionality, functional impairment, and severity of SUD symptoms. Unlike part one, we developed specific hypotheses, based on observations in the preclinical literature, but also informed by the limits of the external measures used in our study. We hypothesized that:

1. Intrusive Thoughts about alcohol use would be negatively associated with indicators of trait impulsivity such as lack of perseverance and lack of premeditation. Further, that Intrusive Thoughts would be related to markers of trait negative emotionality and functional impairment in social and personal domains.
2. Craving would be positively associated with markers of urgency, trait negative emotionality, social and occupational impairment, and impulsive action in real life.
3. Emotionality would be positively associated with markers of urgency and sensation-seeking, trait negative emotionality, social impairment, and impulsive action in real life.
4. Loss of control would be positively associated with lack of perseverance and lack of premeditation, trait negative emotionality, occupational and physical impairment, and would show the highest association with impulsive action in real life.
5. The Compulsive Alcohol Use higher-order factor (CAU) would be positively associated with markers of urgency, trait negative affect, overall functional impairment, and more impulsive action in real life.
6. Higher CAU factor scores would be positively associated with greater severity of AUD symptomatology and would predict AUD symptoms over and above measures of quantity and frequency of use.

### Methods

#### Measures

Descriptive information regarding measures can be found in Table 4. These included the:

**Table 4.**
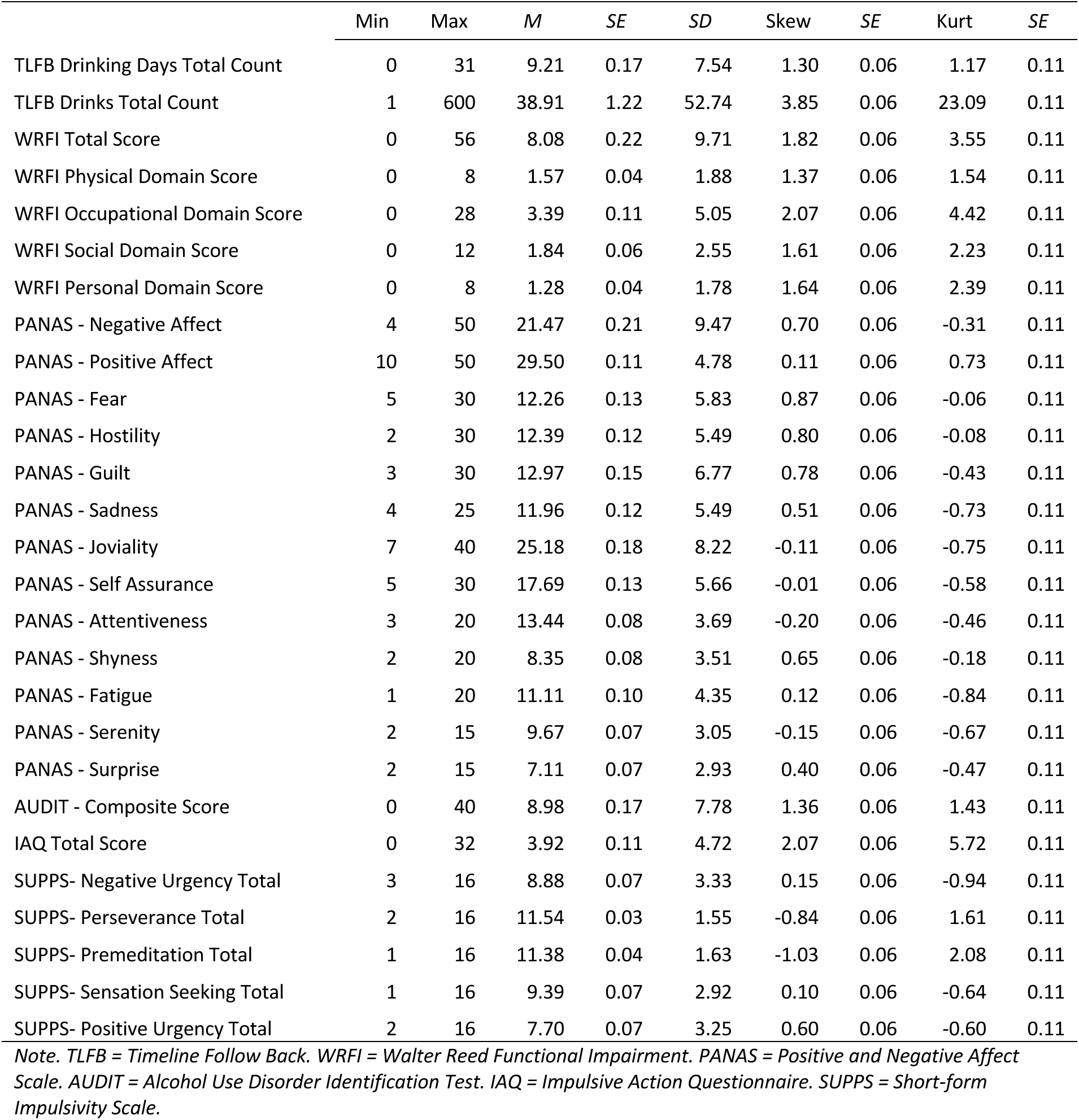
Measure Descriptives.

**Walter Reed Functional Impairment Scale (WRFIS).** The Walter Reed Functional Impairment Scale (WRFIS; Herrell et al., 2014) is a 14-item scale designed to assess current functioning in four areas (physical, occupational, social, and personal) in generally healthy groups. Participants are asked to rate how much difficulty they currently have completing different activities, on a 5-point Likert scale, from *No difficulty at* all to *Extreme difficulty.* Importantly, though this scale was originally designed for use in veterans, the items reflect a relatively high baseline of functioning making it suitable for the present study, with small wording changes. For example, one item asked about “ability to do physical training”, for the present study “physical training” has been replaced with “exercise/physical activity”.

**Timeline Follow-Back (TLFB).** The TimeLine Follow Back Self-Report Procedure (Sobell & Sobell, 1995) is a self-report measure of the quantity and frequency of alcohol use in the past month. Participants are asked to consider their drinking in the past month and complete a calendar indicating their alcohol use on each day, including whether they drank, what they drank, and how much alcohol they consumed. The timeline follow-back has been validated through subdermal alcohol measurement (Simons et al., 2015), and has been validated for both pen and paper and online use (Pedersen et al., 2013).

**Alcohol Use Disorder Identification Test (AUDIT).** The Alcohol Use Disorder Identification Test (AUDIT; Saunders et al., 1993) is a 10-item screening questionnaire assessing alcohol use disorder symptoms and alcohol use consequences on a scale of 0 to 3 (response options change based on question). Items include “How often do you have a drink containing alcohol?” (Answered [0] *never*, [1] *monthly or less,* [2] *2-4 times a month*, [3] *2-3 times a week*) and “Have you or someone else been injured because of your drinking?” (Answered [0] *No* or [2] *Yes, but not in the last year*). We used this measure as a dimensional screener for alcohol use problems.

**SUPPS-P Impulsive Behavior Scale.** The SUPPS-P is a shortened version of the UPPS-P Impulsive Behavior Scale (Whiteside and Lynam, 2001; Cyders et al., 2014) consisting of 20 items. Respondents answer on a four-point Likert-type scale ranging from 1 *strongly agree* to 4 *strongly disagree*, to items such as “I tend to give up easily.” There are four items for each of the five scales (Negative Urgency, Positive Urgency, Sensation-Seeking, Perseverance, and Premeditation). This shortened measure reflects the hierarchical factor structure found in the full-length version; the 2nd order factor of Emotion Based Rash Action consists of Positive and Negative Urgency, Sensation Seeking stands alone, and Deficits in Conscientiousness consists of (lack of) Premeditation and Perseverance.

**Impulsive Action Questionnaire (IAQ).** The Impulsive Action Questionnaire (IAQ; Delawalla et al., In Preparation), is a 32-item measure capturing a variety of specific impulsive actions described in past research. The items capture recent impulsive behavior related to dating and sexual behavior, alcohol use, antisocial behavior, substance use, and change behavior. Participants respond by answering yes or no to having taken the described action in a specific timeframe. For example, “In the past 30 days have you purged after eating a large meal” or “driven recklessly”? Items are summed to provide a raw score reflecting impulsive actions.

**The Positive and Negative Affect Schedule-Expanded Form (PANAS-X).** The PANAS-X (Watson and Clark, 1999) is a 60-item measure assessing affect that has been experienced by the participant “in general.” The PANAS-X consists of two higher order scales assessing general trait positive and negative affect, as well as three facets assessing basic positive (i.e., Joviality, Self-Assurance, Attentiveness) and negative (i.e., Fear, Hostility, Guilt, Sadness) affect, as well as other affective states (i.e., Shyness, Fatigue, Serenity, Surprise). Participants indicate to what extent, on a scale of 1 *very slightly or not at all* to 5 *extremely* they have experienced different feelings, such as “irritable”, “proud”, or “sheepish”.

### Data Analysis Plan

In line with Cronbach and Meehls (1955) suggestions for construct validity, we used regression to examine the nomological network of the derived factors. Specifically, in the full sample, we regressed external measures onto the factor scores derived from the factor analysis to test the discriminant and convergent validity of each compulsive alcohol use model component derived above.

### Results

Descriptive information for external measures can be found in **Table 4**. Given the sample is not clinical and we are assessing a phenomenon that is clinically relevant, we examined AUDIT score frequencies in the harmful to severe range to ensure enough variability. Of the sample, 19% are in the harmful range with scores between 8 and 14 and 18% are in the severe range with scores above 15.

To better understand the core of each factor in the four-factor, higher order model we regressed the factors onto variables of interest assessing impulsivity, emotionality, and functioning. Regression results can be found in Figure 3, (all tabled results can be found in supplemental Materials Table 4). In the proceeding text, we describe and interpret only those pattern of results that are statistically significant.

1. As hypothesized, *Intrusive Thoughts* about alcohol use was negatively associated with lack of perseverance and lack of premeditation (suggesting it is associated with premeditation and perseverance), trait fear, and functional impairment in social domains. Contrary to hypotheses, the *Intrusive Thoughts* was positively associated with trait joviality and self-assurance, negatively associated with surprise, negative affect, and positive affect. Further, it was positively related to occupational impairment.
2. In line with expectations, *Craving* was positively associated with markers of urgency, social impairment, impulsivity in daily life, and trait fear (an indicator of trait negative affect). *Craving* was not associated with occupational impairment or broad trait negative affect, and it was unexpectedly positively associated with trait self-assurance.
3. *Emotionality* results were mostly in line with hypotheses, with some exceptions. Emotionality was not associated with sensation-seeking, was negatively associated with trait negative affect, and showed some unexpected associations, including positive associations, with trait fear and self-assurance and negative associations with attentiveness, fatigue, lack of premeditation, and lack of perseverance.
4. Contrary to hypotheses, *Loss of Control* was *negatively* associated with lack of perseverance and lack of premeditation, and trait negative emotionality. It was not associated with any marker of impairment, and while it was associated with impulsive action in real life, it had the smallest effect size amongst the four factors. There were also unexpected findings—*Loss of Control* was positively associated with trait fear, and self-assurance, and was negatively associated with attentiveness and fatigue. Finally, it was positively associated with urgency.
5. Regarding the Compulsive Alcohol Use higher-order factor (CAU), we hypothesized positive associations with markers of urgency, trait negative affect, overall functional impairment, and impulsive action in real life. Hypotheses were partially supported. CAU was positively associated with urgency, social functional impairment, and impulsive action in real life. It was also positively associated with trait fear, however it was *negatively* associated with negative affect. Likewise results differed from hypotheses in that CAU was negatively associated with lack of perseverance and premeditation, trait attentiveness and trait fatigue and positively associated with self-assurance.

**Figure 3.**
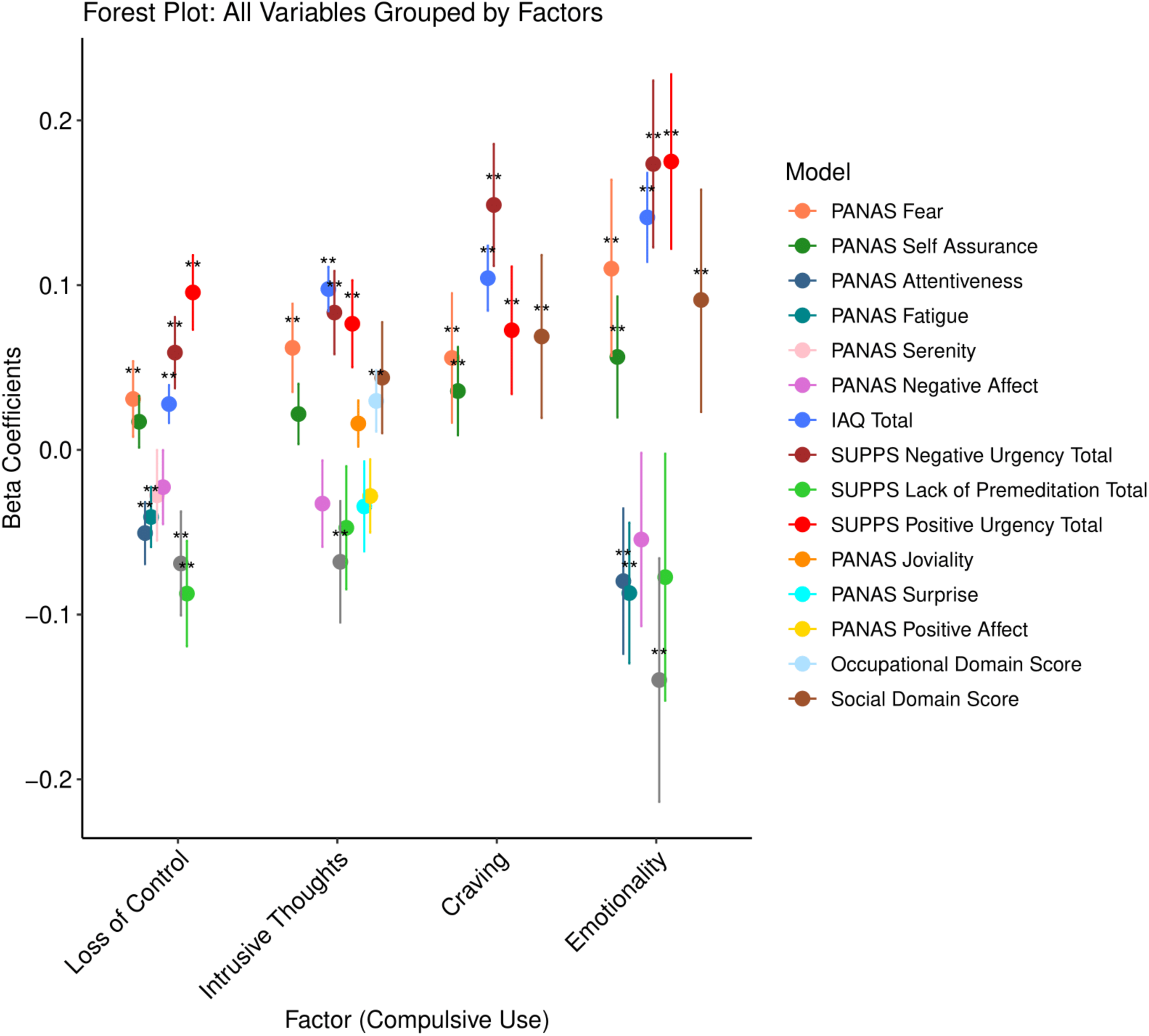
Regression Analysis. Note. Regression coefficients of collateral validation measure with each of the four CAU factors.

To further evaluate the broad construct of Compulsive Alcohol Use, we conducted a hierarchical regression with quantity and frequency of use to predict alcohol use related problems as assessed by the AUDIT (excluding count items). Block one was statistically significant (F(2,1873) = 430.71, *p* <.001, *R^2^* = .32), suggesting both quantity (β= .46, t = 16.91, *p* < .001) and frequency of use (β = .13, t = 4.89, *p* < .001) were associated with alcohol use related problems. The addition of the CAU factor scores (β = .55, t = 32.80, *p* < .001) in block two resulted in significant improvement in prediction of the outcome (F(3,1873) = 810.93, *p* < .001, *R^2^* = .57). Overall, when quantity and frequency were included in the model, they explained 32% of the variance and the inclusion of the CAU factor scores increased the variance explained to 57%. Notably, in block two, frequency was no longer significant in the model (β = .02, *p =* .415) and the variance explained by quantity of use decreased (β = .32, *p* < .001).

## Discussion

In a representative sample, we used EFA and CFA to establish a model of Compulsive Alcohol Use comprising intrusive thoughts, craving, emotionality, and loss of control. The selected model is both theoretically relevant *and* data driven, such that it had good model fit and was associated with external measures that are in line with clinical and pre-clinical literature in the area. Specifically, we found CAU to be associated with emotion driven impulsivity, perseveration and premeditation, social impairment, trait-based fear, energy, and a lack of attentiveness.

In part one, we established the model of compulsive alcohol use. One notable decision point in this process was the inclusion of a fourth factor that captured emotionality related to alcohol consumption. Fit statistics were similar between models with three versus four factors. Further, the three-factor model was more similar to a combination of components that makes up compulsive use in the preclinical literature. However, we suspected that emotionality was not included in preclinical components due to the difficulty of capturing emotion-based motivations for consumption in model organisms (though attempts have been made; Kuo et al., 2022 for review). The distinct content and different patterns of association related to the fourth emotionality factor compared to the craving factor, in the validation stage, suggests the decision to select the four-factor solution was well founded. Additionally, most theoretical models of addiction include some aspect of emotion (e.g., Koob and LeMoal, 2008) and a large body of literature has investigated the role of emotion in addiction (see Stellern et al., 2023 for review).

A second decision point in establishing the CAU model was whether to select a correlated factors or higher order factors model. The correlated factors model had marginally better fit than the higher order factors model, however, one key theoretical consideration was that compulsive use appears to be the sum of its parts. Specifically, use can be labeled compulsive when all four factors are present—otherwise, it is an inherently different pattern of use. For example, an individual may have a high score on the emotionality factor, suggesting they drink when they are experiencing strong positive or negative emotion, but if that drinking is not accompanied by ruminative, intrusive thoughts, craving, and a loss of control, then it may appear in daily life as a glass of wine after a challenging day at work. Likewise, if an individual only scores high in loss of control, perhaps they are at risk for one-off binge drinking episodes, but that pattern of use is not consistent enough to be considered compulsive. Selecting the higher order factor model emphasizes that each factor is a necessary, but insufficient aspect of compulsive alcohol use on its own.

In part two, we provided an initial validation effort for this model. While our validation findings were mixed, it was evident that certain aspects of trait impulsivity were more related to CAU than others. Specifically, urgency was significantly related to all CAU factors and sensation-seeking was not at all associated with CAU factors. Interestingly, lack of perseverance and lack of premeditation were *negatively* associated with some factors—suggesting that CAU is a persistent, ruminative experience, one that is *not* marked by ill-considered action or quitting too early. However, the consistent association with impulsive action in daily life suggests a pattern level of poor decision making; this was consistent with prior work by MacKillop and colleagues (e.g., MacKillop et al., 2011).

Contrary to hypotheses, trait-based tendencies towards positive and negative affect were not as consistently related to the factors and when they were associated, the direction of association was opposite of the hypotheses. Other trait-based tendencies were unexpected and may shed light on these findings and a broader implication of CAU. Taken together, this pattern of association could have two implications. First, it may suggest that individuals who drink compulsively simply do not have trait-based tendencies towards positive and negative emotionality. However, given that smaller facets of positive and negative emotionality were significantly associated with CAU factors in unexpected ways (e.g., fear *and* self-assurance), a broader implication may be that compulsive alcohol use occurs at the intersection of vulnerability for internalizing and externalizing psychopathology. These results suggest that individuals with higher CAU factor scores exhibit transdiagnostic markers of internalizing, such as being ruminative and fearful, as well as externalizing markers such as emotion driven rash action.

Altogether, we were able to develop and begin to validate a model of compulsive alcohol use that partially aligns with empirical findings from the preclinical literature. We found that the four subfactors—intrusive thoughts, craving, emotionality, and loss of control—were highly related to one another but distinguishable. This work suggests each factor is a necessary, but alone, insufficient core component of a collection of behavioral, cognitive, and emotional experiences that comprise compulsive alcohol use. Individuals with both internalizing and externalizing vulnerability may be at the highest risk for this pattern of alcohol use. Finally, compulsive alcohol use factor scores were more informative than quantity and frequency counts in identifying individuals exhibiting features of AUD.

Although this work has several strengths, there are notable weaknesses. First, we were not able to obtain biological samples from the participants to confirm alcohol consumption assessed using the Timeline Follow-Back self-report measure. However, there is evidence that Timeline Follow-Back reports are valid measures of alcohol consumption, as verified by transdermal alcohol assessment (Simons et al., 2015).

There are several relevant future directions in further testing and applying this model. Validation of the model in real daily behavior is necessary to draw parallels with the pre-clinical CSU literature. Likewise, the model should be evaluated in the context of other substances. Basic and applied researchers and clinicians may benefit from the development and validation of a short, easy-to-use measure that adequately captures compulsive alcohol use in general and clinical samples and across all English-speaking demographic groups. Such a measure could be used in imaging and behavioral genetics studies to capture more refined phenotypes of severe alcohol addiction. Accounting for heterogeneity in psychopathology has begun to yield promising results in basic science in other areas (e.g., depression; Tozzi et al., 2024; Drysdale et al., 2017) and has the potential to inform addiction science in a similar manner.

## Data Availability

All data produced in the present study are available upon reasonable request to the authors

